# Drivers of COVID-19 variant wave dynamics: inferring oncoming wave size using global data with genomics

**DOI:** 10.1101/2025.09.16.25335896

**Authors:** S. Molan, N.K. Smith, V. Gandhi, M. Li, C. Colijn, C.L. Murall, J.E. Stockdale

**Affiliations:** Department of Mathematics, Simon Fraser University; Department of Community Health and Epidemiology, Dalhousie University; National Microbiology Laboratory Branch, Public Health Agency of Canada; Department of Mathematics and Statistics, McMaster University; Science Policy Integration Branch, Public Health Agency of Canada

**Author notes:** Corresponding author: 8888 University Drive, Burnaby, BC, V5A 1S6, jessica.

**Keywords:** COVID-19, Prediction, Omicron variants, Genomic data, Random forest

## Abstract

The continued evolution of the SARS-CoV-2 virus drove waves of infection worldwide throughout the pandemic. These evolutionary dynamics posed significant challenges for public health forecasting and, specifically, for predicting the size of COVID-19 waves. In this work we leverage a range of global public data, with a focus on features derived from pathogen genomic sequences, to model and predict the relative size of COVID-19 waves (as compared to the previous wave) across countries. Focusing on Omicron BA.1 and BA.2, we develop statistical models to assess the predictive power of these data in forecasting future variant-driven wave peaks. We find that, while forecasting wave size is a challenging task, variables such as genomic variant characteristics, prior wave dynamics, and demographic features e.g. life expectancy were informative, whereas seasonality was not. Our results show that the importance of features changed markedly between Omicron waves, reflecting the evolving epidemiological and genomic landscape. This work provides insights into improving predictive models for future outbreaks and pandemics, and prioritizing data collection efforts to enhance forecasting accuracy.

## 1 Introduction

Since the appearance of SARS-CoV-2 in late 2019, heterogeneity in public health response, vaccine uptake, health care system capacity, and the continued evolution of more transmissible and immune-evasive variants created variable transmission dynamics, with multiple waves of COVID-19 cases per year in most countries. Many respiratory viruses have clear seasonal wave dynamics, with increased transmission and activity during the colder months, but the wave dynamics of COVID-19 appear to have had faster persistent oscillations. COVID-19 waves have been less predictable in timing or magnitude, posing a challenge for public health preparation and response.

During the pandemic, surveillance and data collection efforts made unprecedented amounts of global public data available, including epidemiological, genomic, clinical, serological, mobility, and demographic data. There were many efforts to collect and standardize population-level data for public use. For example, the COVID-19 Data Repository by the Center for Systems Science and Engineering (CSSE) at Johns Hopkins University collected basic epidemiological data such as reported cases, hospitalizations and mortality from all countries around the world^1^; similarly the World Health Organization (WHO) collated global epidemiological data^2^; Apple and Google mobility released population-level aggregated movement data publicly to support COVID-19 research^3, 4^; GISAID hosted SARS-CoV-2 genomic sequences^5^; SeroTracker collated the results of sero studies globally^6^; the Oxford COVID-19 Government Response Tracker reported on the changing levels of government pandemic policy and interventions^7^; and Our World in Data (OWID)^8^ collated and shared data from many of the above sources. Compared to any historical disease outbreak, there is a very large volume of publicly available data for COVID-19.

Despite the dissimilarities of wave dynamics between countries, the emergence of the Omicron variant (B.1.1.529) in late 2021 was relatively consistent around the world. In a span of approximately three months, the Omicron wave overwhelmed the testing and epidemiological data collection capacity of many jurisdictions. Due to its significantly higher rate of transmission and strong immune evasive properties, Omicron and its descendants completely replaced the previous variants that were more genetically similar to the wildtype. Omicron continued to evolve immune evasive variants with very successful lineages that circulated year-round. With hundreds of circulating lineages at any given time, monitoring and identifying the next fittest variant that would drive an uptick in cases or evade currently used drugs or vaccines became a collective non-stop effort by scientists around the world^9–11^. National and local public health authorities monitored the activity of COVID-19 in their jurisdictions and compared to other countries (e.g. neighbouring countries or countries that had a head start in the emergence of the latest variant of interest) to evaluate the magnitude of the impact of the next wave. These wave dynamic comparisons were particularly important at the height of the pandemic, as decisions on whether to implement or release public health measures were responsive and critical. However, despite the large volumes of real-time globally shared data, this predictive work was done largely using human intuition that tried to synthesize a myriad of likely predictive factors (current season, differences in population vaccination status, timing of previous wave, etc.) along with the various variant characteristics and current epidemiological trends and thus, understandably, fell short of reliably predicting important indicators of impact, such as the relative size of the next local wave.

In this work, we collate a wide range of publicly available data, including epidemiological, genomic, clinical, serological, mobility, and demographic data, to explore the wave dynamics of Omicron BA.1 and BA.2 in a variety of countries. In particular, we focus on the inclusion of features derived from global SARS-CoV-2 genomic sequences. We seek to determine whether the relative size of an upcoming case peak (BA.1 or BA.2, compared to a recent peak) could be predicted using a statistical model and data up until the initial growth phase of the respective variant. We also seek to identify which of the features were the most informative of the relative wave peak sizes, with a focus on the genomic features derived from global sequence data. Alongside this manuscript, we make available our curated and cleaned dataset which collates this wide range of features, from December 2019 until January 2023, across 248 countries and jurisdictions.

## 2 Materials and Methods

### 2.1 Prediction task

We focus on predicting the peak size of a new variant-driven wave relative to the peak size of the previous wave in a country. We study two waves: Omicron BA.1 relative to Delta and Omicron BA.2 relative to BA.1, which we refer to as BA.1/Delta and BA.2/BA.1, respectively. These periods corresponded to a time of major change in the pandemic, with pronounced increases in transmission. Epidemiological and genomic data sources were still regularly collected at this time, along with data on COVID-19 testing and vaccination/booster campaigns. In the prediction task, our response variable is the relative size of the maximum case counts attributed to the emerging variant of concern (VOC) vs. the maximum VOC-attributed case counts in the previous wave (a different dominant variant). For example, the relative size of BA.2 is the ratio of the peak cases of BA.2 and of BA.1. Compared to the task of predicting absolute peak sizes, this minimizes the impact of variation in the levels of case detection in different countries. Although case count time series are impacted by changes in at-home test reporting, access to testing and so on, it remains the case that reported infections were one of the primary ways countries assessed their COVID-19 burden, and of high interest for public health decision makers. We seek to assess how routinely collected data, particularly genomic sequence data, could serve as an early indicator of the relative wave size, for which case counts are the earliest measure.

In the sections below, we first describe the full collated dataset, for all available countries from the start of the pandemic until the end of January 2023. We then describe the subset of the data that is used in our predictive model.

### 2.2 Data

We collated publicly available data on variables that could be expected to affect the size of COVID-19 waves across countries. Data were obtained from online repositories including Our World in Data (OWID)^8^, which collected information from sources such as the Johns Hopkins University COVID-19 data repository^1^, the WHO database^2^, and the Oxford COVID-19 Government Response Tracker^7^. Additional data came from SeroTracker^6^, Google Mobility^3^, and GISAID^5^. Some variables were taken directly from these sources (e.g., case counts), and some we derived from the available data, as will be described in the data processing section. Where available, we collected a daily time series of each variable for each country (a total of 248 countries and jurisdictions) throughout a study window of 2019-12-29 to 2023-01-29.

We sourced global viral genomic data of human origin (not animal or environmental) from GISAID^5^ on May 4, 2023. Sequences were filtered to include only those that had complete collection dates and were submitted on or before February 1, 2023. The variants were designated using Pangolin version 4.2^12^, PUSHER-v1.18.1.1. We then classified Pango lineages into variants of concern (VOC) and variants of interest (VOI) groups that align with the Nextclade clade classification system (a higher-order grouping of successful lineages^13, 14^). Full details on the VOC and VOI classifier function are available in the GitHub repository associated with this work, as is a supplemental table describing the 1,048,516 sequences used, available on GISAID and accessible at doi.org/10.55876/gis8.241023pk. To infer the dates of global emergence of each clade, a method that uses the time to the most recent common ancestor (tMRCA)^15^ was applied to each Nextstrain clade. A curated date for Omicron BA.1 was taken from Viana et al.^16^. To avoid sequences with data entry errors, we removed any sequences dated prior to the inferred date of global emergence of that particular lineage’s clade. We collapsed the genomic metadata dataset into a weekly format from Monday to Sunday, with the week’s total counts being summed on Sunday. To obtain mutation and amino acid substitution calls for each sequence, the global viral sequence dataset was run through Nextclade CLI^17, 18^ v. 3.4.0 that aligns the genomes relative to the wildtype (Wuhan-Hu-1/2019 (MN908947)), assigns clades, runs quality checks and gives mutational information relative to the original Wuhan reference. To minimize noise in the data for the predictive modelling task, upon combining genomic data, we converted all data to a weekly timescale, in which weeks run from Monday to Sunday.

In total, we collated data on 113 variables, which we group into 11 broad categories: COVID-19 cases and reproduction rates, deaths, excess mortality, hospitalizations, policy responses, testing, vaccination, serology studies, genomic variables, demographic, and miscellaneous. The final category includes variables such as mobility data, season, and latitude/longitude. The full list of variables is shown in Figure **??**. The full dataset, along with variable descriptions and sources is available in the GitHub repository associated with this work.

These data draw from many different sources and are subject to many limitations, including missing data (almost certainly not at random), changes in data collection policy over time, differences in definitions across countries, and more. Despite this, these data represent perhaps our most comprehensive international understanding of the burden of COVID-19, the immunity against it, and the viral evolution of SARS-CoV-2 that is publicly available, so, noting these limitations, we proceed.

### 2.3 Data processing

To summarize the genomic data in a variant-specific way, we derived additional variables, including VOC-specific case counts, a measure of sequencing effort, genetic diversity and distance, and growth advantage.

We estimated the case counts for each VOC by multiplying the total weekly case counts (from OWID^8^) by the proportion of the total VOC sequences (from GISAID^5^). This generated a time series of weekly case counts per VOC per country. During the emergence of each new variant, we calculated the time (number of days) since the previous variant’s peak. We created a sequencing effort variable to capture the variation in case testing and genomic sequencing between and within countries throughout the pandemic. We defined sequencing effort as the total weekly number of sequences divided by the total weekly number of cases in each country.

For each country, we calculated several diversity metrics. The simplest was richness, defined as the total number of unique Pango lineages detected per week. We also included the number of distinct VOCs present as a separate variable. To account for the relative abundance of lineages, we calculated Hill diversity of order 1 (*D*_1_), equivalent to the exponentiated Shannon index (*H*), which incorporates both lineage richness and evenness, providing a more complete picture of diversity than richness.^19^. The Shannon index was calculated using the *vegan* package^20^ with the formula:

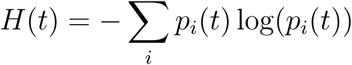

where *p_i_*(*t*) is the proportion of sequences belonging to the Pango lineage *i* at time *t*. The Hill diversity was then calculated as *D*_1_(*t*) = exp(*H*(*t*)).

To quantify the genetic distance between an emerging variant and concurrently circulating lineages, we calculated the mean number of unique amino acid substitutions in the circulating lineages relative to the emerging variant (Supplementary A.1). For each emerging variant, a high-quality and representative early reference sequence was manually selected. For example, the BA1 reference sequence was from Botswana on 2021-12-20 (GISAID Accession ID: EPI ISL 9002788), and the BA.2 reference sequence was from Denmark on 2021-12-20 (GISAID Accession ID: EPI ISL 8347448). Within each country’s respective fitting window (see below), we computed the genetic distance between the emerging variant’s reference and every sequence in the dataset. The mean of these distances was then calculated for both the S-gene and the total genome, which were used as features in the model. Further details can be found in the Supplementary Materials.

To include the relative fitness of the variants, we estimated the selection coefficient (*s_i_*) for each emerging variant. We fitted a multinomial logistic growth model to variant proportions in each country during defined fitting windows. These selection coefficients have become the gold standard for estimating the relative growth rate of emerging variants (e.g. CoV-Spectrum.org^21^) and are used to quantify the speed of variant replacement; faster-growing variants are considered more likely to drive subsequent waves of infection. Here, the frequency *p_i_*(*t*) of variant *i* at time *t* is expected to increase relative to all other circulating variants by

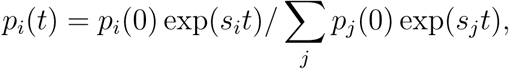

where *s_i_* is the selection coefficient of variant *i* per day, assuming constant selection for the fitting period. We followed the fitting procedure and the code base developed by members of CAMEO (CoVaRR-Net’s Computational Analysis, Modelling and Evolutionary Outcomes team^22^), which is used in the Canadian real-time variant tracking site *Duotang*^23^ and is described by Gill et al.^24^.

Serology data was available as a single numeric value per serology study, rather than a time series. To handle this, we used Last Observation Carried Forward (LOCF) imputation to generate a piecewise constant time series. This carries forward the percent seroprevalence identified in each study from the start of that study’s time window, through to the start of the next seroprevalence study (or until February 1 2023, whichever is earlier). We selected LOCF for its simplicity, as it avoids making assumptions such as linear growth in seroprevalence in the unobserved periods between studies. We performed a similar LOCF imputation for the vaccine manufacturer data, which shows how many of a given manufacturer’s vaccines were used in a given country. No further adjustments or corrections were made to the raw data at this stage.

### 2.4 Modelling approach

While our master dataset covers the entire pandemic up to February 2023, for the task of predicting BA.1/Delta and BA.2/BA.1, we focused on the subset of the data corresponding to the time period of emergence of the respective new variant. This was designed to model the time in which the question of emerging variant wave size would be of most interest to local authorities. We mask from the predictive model the knowledge of any data after the end of a fixed fitting window.

We defined the fitting window as the week in which the emerging variant passed 10% of weekly sequences in the country, and the four weeks prior (35 days total). For example, if the emerging variant passed 10% in frequency on 2022-02-13, then the fitting window ranges from 2022-01-10 to 2022-02-13. This means that each country has its own fitting window for each wave, relative to the variant’s growth in that country. We label *t*_0_ as the first Sunday on which the emerging variant’s sequences passed 10% of the total. The previous 4 Sundays are labelled *t_−_*_1_, *t_−_*_2_, *t_−_*_3_, and *t_−_*_4_, resulting in a fitting window of [*t_−_*_4_, *t*_0_]. We trained our model on slices of the master data set within the fitting window (that is, all the features from dates *t_−_*_4_, *t_−_*_3_, *t_−_*_2_, *t_−_*_1_, and *t*_0_). An imbalance in the total number of variables (113) compared to the number of countries with sufficient data for prediction (below) motivated the use of multiple data points per country. To further minimize this imbalance, we discarded some redundant variables, prioritizing (i) summary variables and (ii) those with least missingness. For example, we discarded daily new tests per 100,000 population and individual policy measures such as school closures, but retained daily new tests and overall stringency index.

Many countries had significant amounts of missing epidemiological data or relatively limited genomic data. To ensure our analysis focused on countries with robust genomic surveillance, we selected a subset of countries based on their sequencing effort. We identified 43 countries (Table **??**) that maintained a minimum of 10 sequences per week between 2021-06-01 and 2022-06-01. These countries were also found to have the most robust data across other variables. The subset of 25 countries belonging to the Organization for Economic Co-operation and Development (OECD)^25^ (Table **??**) had further decreased missingness: the results of a model trained on these OECD countries are included in the Supplementary Materials. In the rest of the main text, we focus on a model trained on 46 variables (Table **??**) and 215 (43 *×* 5) observations for 43 countries.

We used multiple imputation to handle the remaining missing data, using the *mice*^26^ package in R with Predictive Mean Matching (PMM), excluding the response variable of relative peak size. We observed, across all countries, an all-or-nothing pattern of variable reporting: for each country and variable, data were either fully reported across all five time points or entirely missing. Therefore, we considered the “country” variable as a blocking factor for the imputation. We performed 30 separate imputations for each missing value, generating a set of plausible datasets to take into account imputation uncertainty.

The Random Forest algorithm was chosen for its ability to model complex interactions among variables. For each imputation of the dataset, the *randomForest*^27^ package in R was used to train a random forest model on a training set. We implemented a leave-one-out cross-validation (LOOCV) procedure. For each iteration, all five data points from one country were held out as the test set, while the remaining countries were used for training. We used grid search to identify the optimal value for the number of features tried at each split, and each random forest model was configured to grow 100,000 trees. Convergence was monitored through the Out-Of-Bag (OOB) error rate, ensuring that the model’s error rate stabilized as the number of trees increased.

Predictions were made for the held-out country using each of the 30 imputed dataset models. These were averaged to obtain a single prediction per observation. The average predictions were compared against the ground truth relative wave sizes to compute the Root Mean Squared Error (RMSE) and the Coefficient of Determination *R*^2^. Additionally, the model’s ability to correctly predict whether the subsequent wave is larger or smaller than the last one was assessed by classifying predictions into four quadrants based on the true and predicted ratios, with correct classifications falling into the first and third quadrants. Lastly, feature importance was calculated for each imputed dataset and aggregated to understand the overall importance of each feature in predicting the target variable.

## 3 Results

Figure 1 shows the overall VOC dynamics from 2020 to February 2023, derived from total global case counts (top) and the VOC proportions from global whole genome sequencing data (top and middle). The bottom panel shows the VOC replacement wave dynamics, illustrated by the VOC-specific case waves. Note that, with the exception of Alpha and BQ, all VOCs had reached *>* 85% of the total global weekly cases (middle panel, Figure 1) by the time case counts peaked globally (top panel), demonstrating a tight temporal coupling between the VOC replacement dynamics and the case wave dynamics. In the first year of the pandemic, wildtype variants dominated and peaked in late 2020. The emergence of the VOCs in early 2021 led to more oscillatory dynamics, with Al-pha and Delta dominating during 2021. However, the emergence of Omicron was a massive global contagion event that led to the largest infectious disease wave ever recorded. Following the prominent BA.1 wave was a global smaller BA.2 wave, though case counting had been overwhelmed in many countries after BA.1, and thus all Omicron waves should be considered under counted, particularly beyond 2023 as public health policies shifted away from COVID-19 case counting.

**Figure 1:**
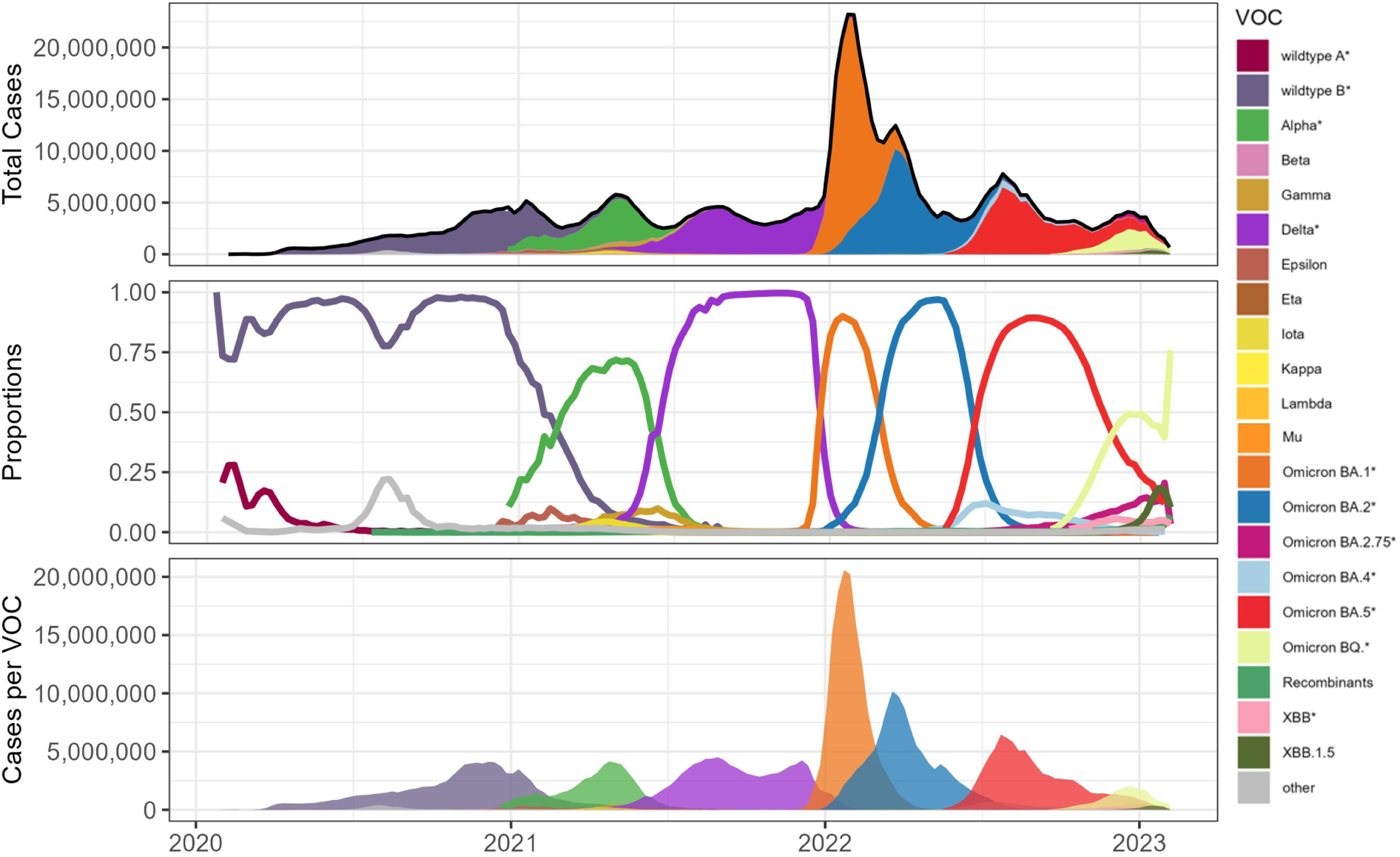
World VOC dynamics. Top panel: Total global reported cases filled in with stacked VOC proportions (middle panel). Middle panel: global VOC proportion plot from sequencing data. Bottom panel: VOC-specific cases over time (determined by multiplying the total cases by the VOC proportion). VOCs with * includes all sublineages.

While the aggregated global dynamics show a large BA.1 wave followed by a shorter and wider BA.2 wave, the country-level reality varied significantly. In fact, the BA.1 and BA.2 Omicron lineages emerged very close together in time, and whether BA.2 would drive a wave locally was hard to interpret at the time. Figure 2 illustrates this variability among countries. The BA.1 and BA.2 wave dynamics can be divided into four categories: larger BA.2 than BA.1 (e.g. Thailand), equal BA.2 (e.g. Sweden), smaller BA.2 (e.g. Canada), and no BA.2 wave (e.g. Peru). Delta was consistently before the emergence of BA.1, but relative sizes at the country level also varied, despite the picture of a smaller Delta wave depicted by global dynamics in Figure 1. See Supplementary Figure **??** for the full Delta/BA.1 wave dynamics.

**Figure 2:**
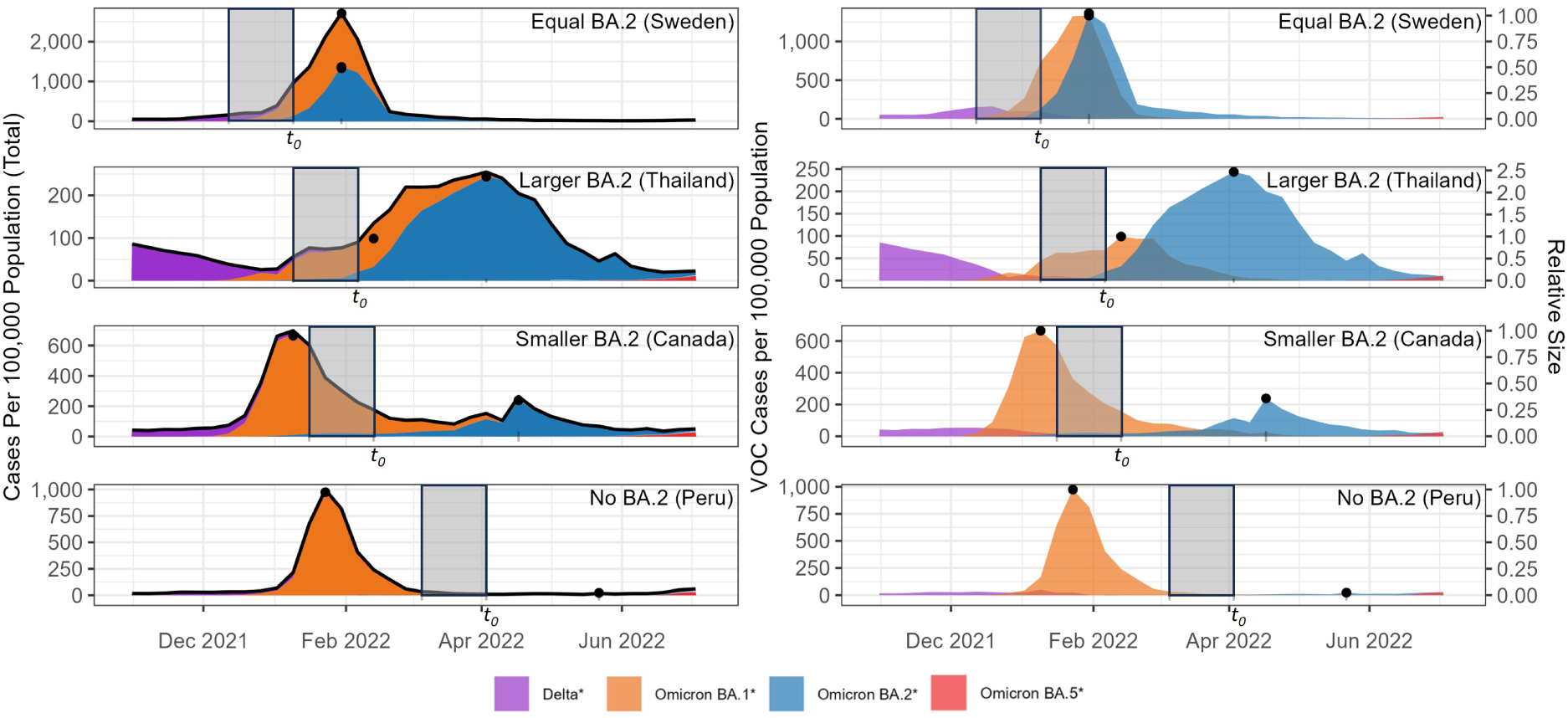
Example categories of country-specific wave dynamics of BA.1 and BA.2. The left column shows the total cases per country filled in by VOC proportion. The right column shows case waves attributed to specific VOCs in a single country over time. The top row shows an example (Sweden) with approximately equal BA.1 and BA.2 waves. The second row shows an example (Thailand) of a larger BA.2 wave. The third row shows an example (Canada) of a smaller BA.2 wave. The bottom row shows the only example (Peru) among the 43 countries with no BA.2 wave. The shaded area represents the five-week fitting window used in our analysis, indexed by the week in which BA.2 exceeds 10% of local sequences (*t*_0_). Black dots represent the BA.1 and BA.2 peaks determined from the VOC-specific cases. In all countries, BA.1 always comes before BA.2, except in Sweden where they peak at the same time. The secondary Y-axis in the right column shows the response in our predictive model, i.e. the relative size of BA.2/BA.1 per country.

In countries with a larger BA.2 wave (defined as relative size *>* 1.1), *t*_0_ occurred at the same time or before the BA.1 peak (Austria, Denmark, Germany, Malaysia, South Korea, India, Singapore, and Thailand, as shown in Figure 2). In countries where BA.2 and BA.1 had comparable sizes (relative size *≥* 0.9 and *≤* 1.1), *t*_0_ occurred before or after the peak of BA.1 (Sweden is before, Finland is after). Of the 32 countries with smaller BA.2, 26 (81%) had *t*_0_ after the BA.1 peak (relative size *≥* 0.05 and *<* 0.9; Argentina, Australia, Brazil, Bulgaria, Canada, Chile, Colombia, Costa Rica, Croatia, France, Ireland, Israel, Italy, Japan, Luxembourg, Mexico, Panama, Poland, Portugal, Slovakia, Slovenia, South Africa, Spain, Switzerland, USA, United Kingdom). In one country *t*_0_ occurred at the same time as the BA.1 peak (Belgium), and in the remaining five (16%) it occurred before the BA.1 peak (Indonesia, Netherlands, Norway, Romania, Russia). In the only country with no BA.2 wave (relative size *<* 0.05), *t*_0_ occurred after the BA.1 peak (Peru). Overall, we see that our fitting windows generally end well before the peak of the wave to be predicted, and indeed often before or during the peak of the prior wave. See Supplementary Figure **??** for the full BA.1/BA.2 wave dynamics.

Selecting a subset of countries to use in the predictive model significantly reduces missing data (Figure 3). The 43 chosen countries had at most 35/113 (31%) entirely missing variables, before data sub-setting and imputation. We were also able to maintain reasonable global representation among our chosen countries. Further visualizations and examples of data and missingness are available in the Supplementary Materials, Figures **??** through **??**.

**Figure 3:**
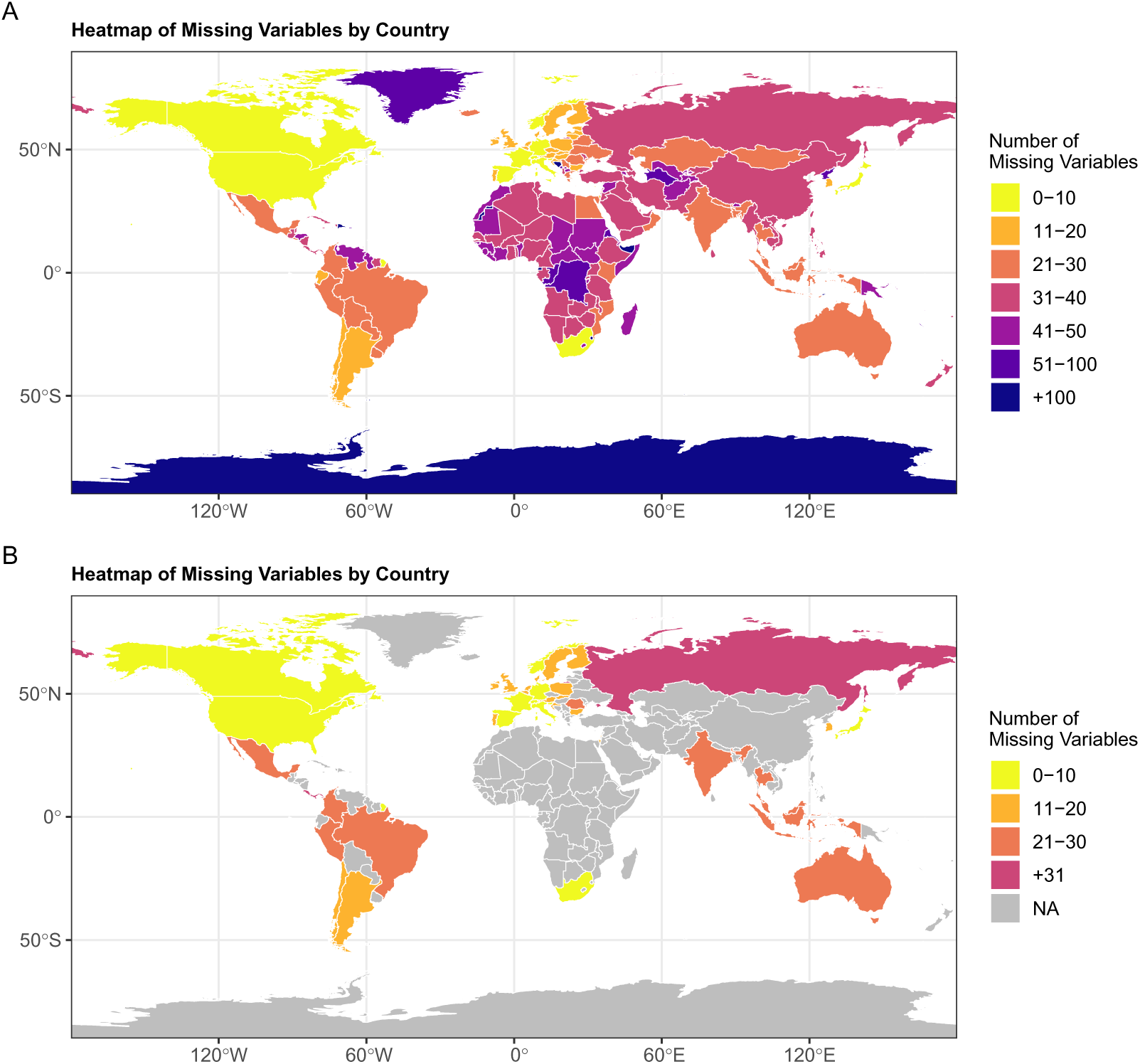
Number of missing variables (out of a total 113) per country, (A) among all countries for which data was collected, (B) among the 43 countries selected for prediction. We define here missing as entirely missing, for all weeks throughout the entire period of study 2019-12-29 to 2023-01-29.

Panel A of Figure 4 shows the predictions of the random forest model, with the response variable modelling the relative size of BA.1 to the Delta wave. The RMSE was 4.09 and *R*^2^ was 0.16. To put this into perspective, the range of observed ratios of BA.1 to Delta was from 0.12 to 34.10.

**Figure 4:**
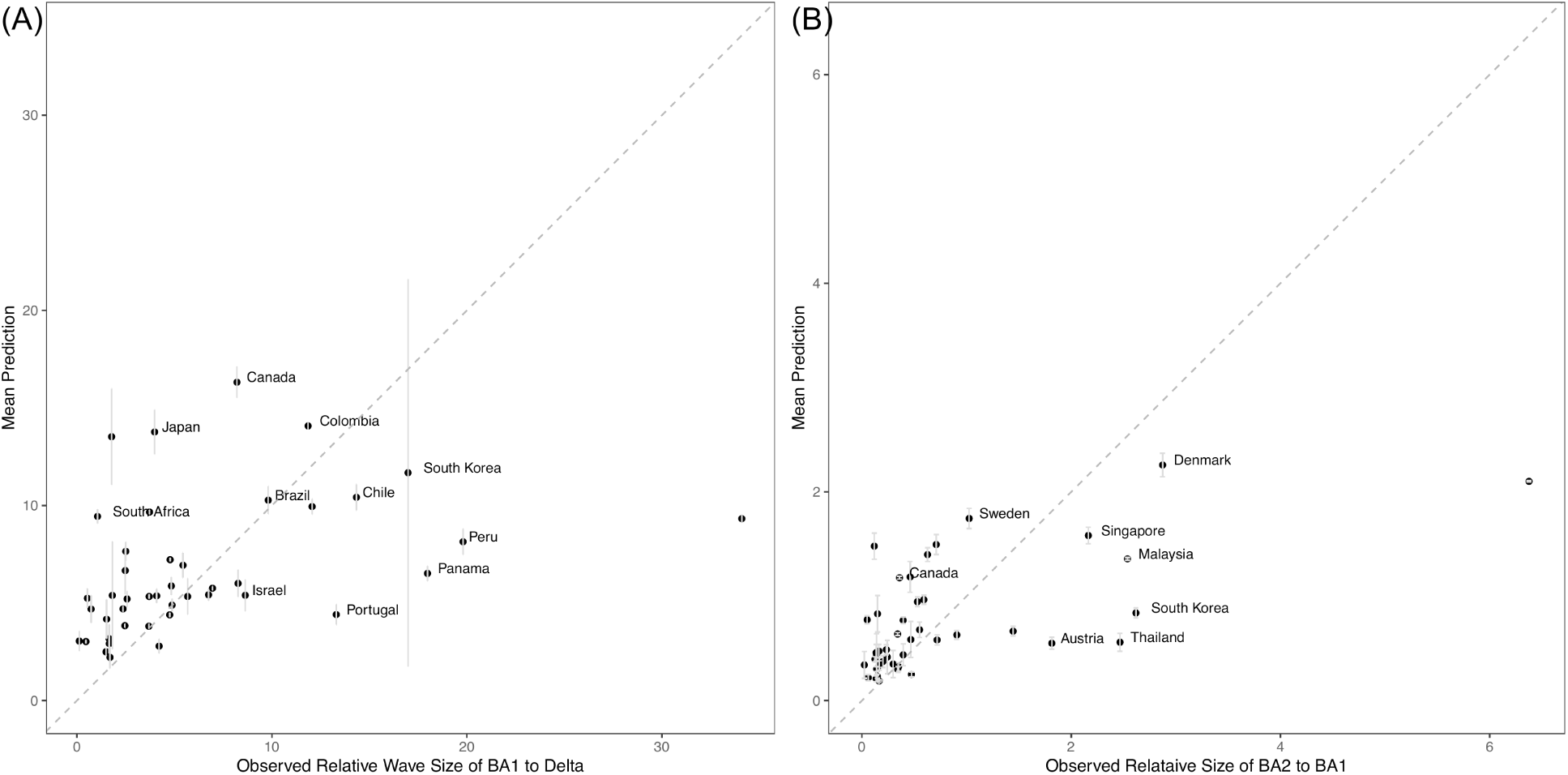
Mean predictions of the wave size of (A) BA.1 relative to Delta, and (B) BA.2 relative to BA.1, across 43 countries using a Leave-One-Country-Out cross-validation approach with a Random Forest model. The x-axis represents the observed relative wave sizes. The y-axis shows the corresponding mean predicted values. The dashed line represents the 1:1 line, indicating perfect agreement between observed and predicted values. Error bars represent the standard deviation of the predictions across multiple imputed datasets.

The prediction for South Korea has a large uncertainty interval. Australia is an outlier. Prior to the BA.1 wave, Australia had an incredibly low number of COVID-19 cases, under their pursued ‘COVID-zero’ strategy. This strategy was ended in late 2021, close to the time of BA.1 emergence globally. Although our model predicted a large growth of cases in Australia (predicted ratio 9.33), it was unable to predict the true scale of such a change (actual ratio 34.10).

Panel B of Figure 4 shows the predictions for the relative wave size of BA.2 to BA.1. The RMSE was 0.57 and *R*^2^ was 0.39. The range of the observed ratios of BA.2 to BA.1 was 0.02 to 6.38. The RMSE of the BA.2/BA.1 model was much smaller than that of the BA.1/Delta model, which is expected given the smaller range of observed ratios for BA.2 to BA.1. India is the outlier here, with a much larger BA.2 wave than predicted. Interestingly, India, along with Denmark and Sweden, had the highest mean genetic distance from the emerging variant BA.2, while also being one of the first countries to detect this lineage (Figure **??** and Figure **??**).

We also calculated how many times the random forest models predicted the general trend of the subsequent wave, i.e., is the next wave larger, smaller, or equal in size to the last wave. This measure of accuracy is perhaps more relevant in practice if the question of public health interest is whether the next wave will be better or worse than the previous one. Our models correctly predicted whether the subsequent wave was larger, smaller, or equal size in 91% and 80% of the countries for BA.1/Delta and BA.2/BA.1, respectively (Figure 5).

**Figure 5:**
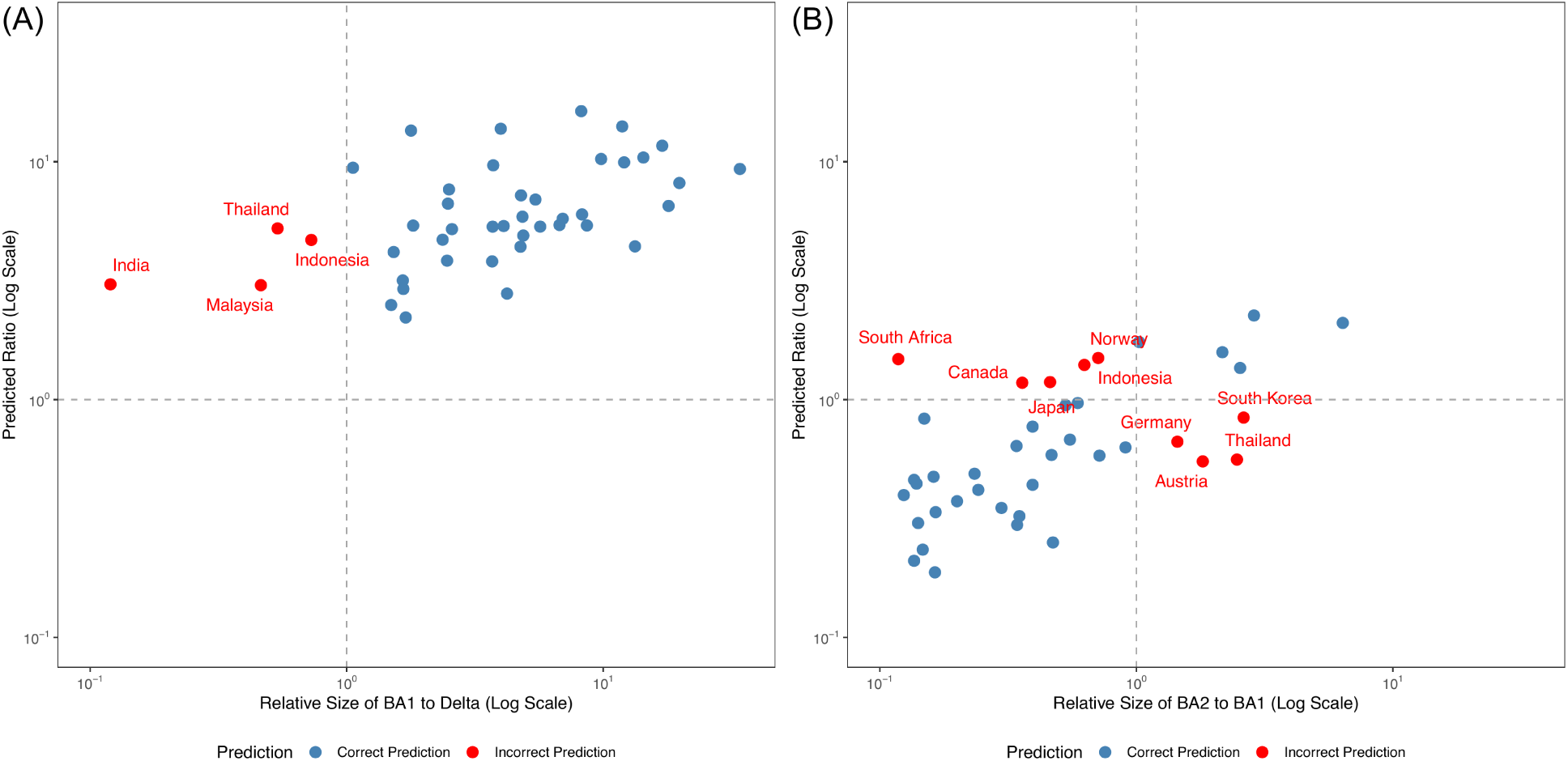
Prediction accuracy plots for two models: (A) BA.1 to Delta and (B) BA.2 to BA.1 variants, both on a log scale. Points in the first (top-right) and third (bottom-left) quadrants represent correct predictions where the model accurately identifies if the ratio of the subsequent waves is larger or smaller than 1. Points in the second and fourth quadrants represent incorrect predictions, where the model incorrectly identifies the ratio of the subsequent waves.

Figure 6 shows the feature importance plots for the two models, visualizing the importance of each predictor variable in contributing to the model’s accuracy. The top panel ranks the features according to the importance score, calculated as the mean increase in the mean squared error (MSE) for the BA.2/BA.1 model when that feature is removed from the model. The bottom panel shows the equivalent scores for the BA.1/Delta model. Genomically derived variables are among the features of high importance. For the transition from BA.1 to BA.2, the genomic distance measure of all genes was the most important feature, although it was only moderately important for the transition from Delta to Omicron. The genomic distance of the S-gene and the selection coefficient were also moderately to highly informative in both models, as was the number of days since the previous VOC peak. We identified several demographic and health variables of importance in each wave, for example, cardiovascular death rates, life expectancy, and longitude were features of highest importance for BA.1/Delta and somewhat lower importance for BA.2/BA.1. Diabetes prevalence was more important in the transition from BA.1 to BA.2. Interestingly, we see considerable changes in feature importance between waves, although several features (e.g., total deaths per million, population density, GDP per capita) are informative across both waves.

**Figure 6:**
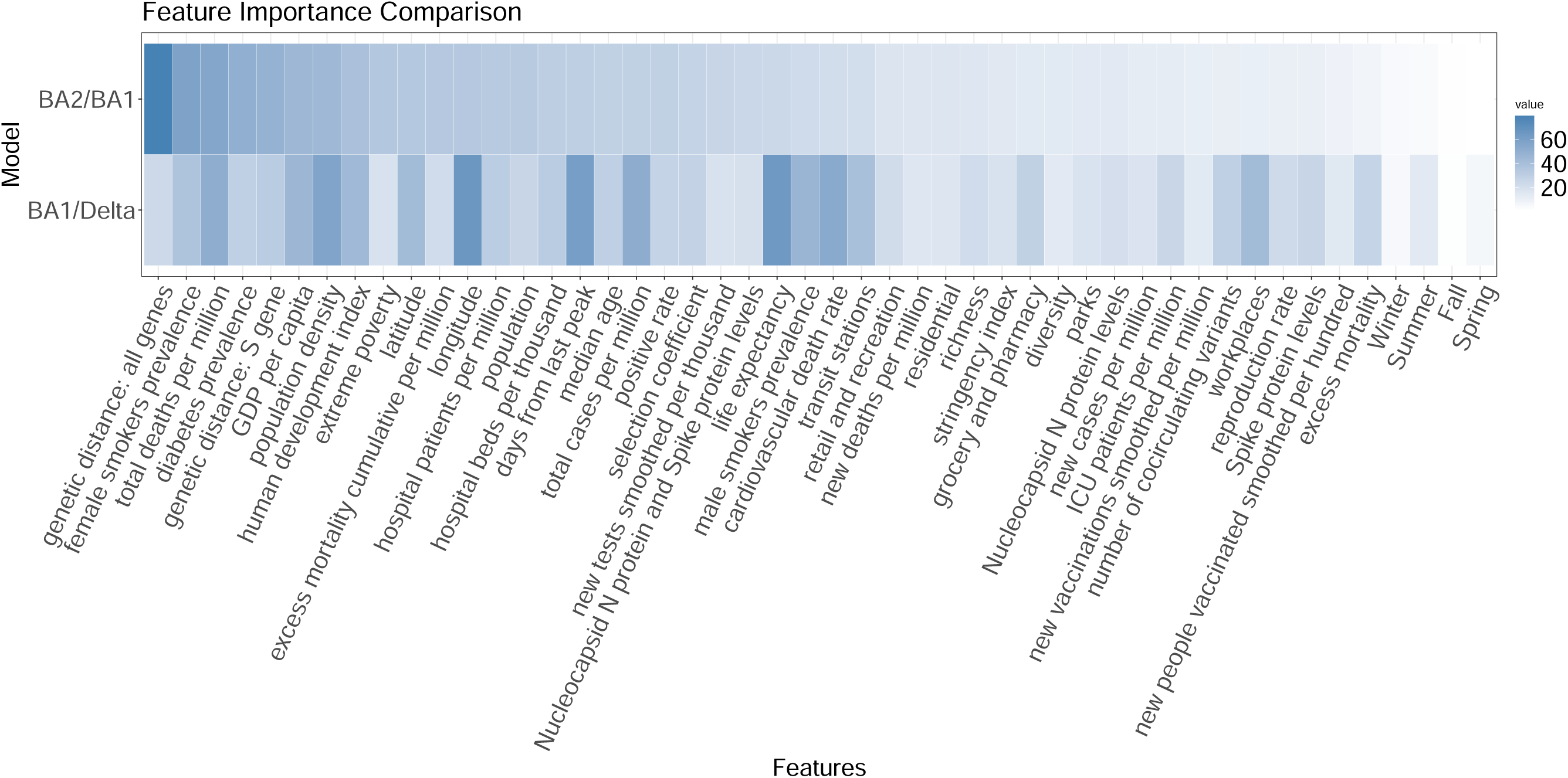
Feature importance comparison for Random Forest models predicting the relative wave size of BA.1 to Delta and BA.2 to BA.1 among 43 countries. The heatmap shows the Mean Decrease in Accuracy (MDA) of various features for each model, with darker shades indicating higher importance.

The stringency index was not found to be important in predicting the relative wave size for either wave, nor were counts of new deaths or cases during the fitting window (although total cases were informative), nor mobility features (parks, residential, etc.) or serology features. However, we note that the missingness in the serology data was high. Of note, all the features pertaining to seasons of the year fell last or nearly last in importance, suggestive that the season a country is in at the time of emergence may not have played a significant role in relative Omicron wave size (a marked departure from what is expected of respiratory viruses).

The results of the 25 OECD countries model are available in the Supplementary Materials. Despite more consistent data quality in the OECD countries, the results were similar. The model trained on 25 OECD countries has slightly higher accuracy in predicting if the next wave will be larger or smaller (100% for BA.1/Delta and 88% for BA.2/BA.1 compared to 91% and 80%, respectively), but RMSE and *R*^2^ are very similar. Notably, all countries had a larger BA.1 wave than a Delta wave. This is a promising result: suggesting that including countries with less consistent data is not detrimental to model performance.

## 4 Discussion

Using a large well-curated global dataset that combines various data sources (epidemiological, genomic, and other data), we find that random forest models can reasonably predict the relative size of a wave (compared to the previous wave). In practice, the most pressing question upon the emergence of a new variant is whether it is expected to have a stronger or weaker impact than the previous variant, and the relative case wave size is seen as an important indicator of local impact: in answering this, our model has very strong performance (Figure 5).

We find that the most important features for the transition between Delta and BA.1 included geographic factors (longitude), population health metrics (life expectancy and cardiovascular death rate), and the number of days since the peak of the Delta wave. This suggests that demographic conditions were the most critical in explaining the shift from Delta to BA.1. During the Delta period, higher-income nations were vaccinated at very high rates, while many lower-income countries struggled to secure vaccines^28^: it is possible that demographic features, such as life expectancy, may capture a broader economic effect on relative wave size through impacts on population immunity.

For the transition between BA.1 and BA.2, the most important features included genetic distance measures (total number of amino acid substitutions and the number of S-gene amino acid substitutions) between the emerging variant and circulating variants, and some population health metrics (total female smokers, total deaths per million, and diabetes prevalence). Note that the total genetic distance was more informative than the VOC’s genetic distance in the S-gene alone. This finding underscores that viral fitness is a polygenic trait and that mutations outside the Spike protein, perhaps influencing replication efficiency or innate immune antagonism, were also critical determinants of Omicron’s success. The association with diabetes prevalence, a known risk factor for severe COVID-19^29^, may indicate that in a population with high levels of baseline immunity from recent infection, the underlying comorbidities become more significant predictors of a variant’s success by marking the remaining pockets of vulnerability. This suggests that both the inherent characteristics of the variants themselves and the demographics were strongly associated with this transition.

We propose that the absence of shared top features between the two waves likely reflects the rapidly evolving host-pathogen landscape. The transition from Delta to BA.1 occurred when global vaccine coverage was highly unequal, making broad demographic and economic factors and the resulting differences in population immunity the dominant predictors. In contrast, the subsequent transition from BA.1 to BA.2 occurred after the massive BA.1 wave had generated more widespread (although short-lived) immunity, likely increasing the relative importance of the subtler genetic differences between these two closely related sublineages. This finding suggests a challenge for pandemic forecasting more broadly: if driving factors are markedly changing dynamically, we cannot naively rely on models trained on previous waves to predict future waves.

Although seasonality was considered in the analysis, it did not show a strong influence on variant dynamics in either wave studied. This finding challenges the assumption that COVID-19 would evolve into a purely seasonal virus. The absence of seasonality as an important feature has implications for public health, suggesting that COVID-19 waves could occur outside of typical respiratory virus seasons, affecting clinical capacities and complicating vaccine roll-out plans, as seen with recent summer waves. The rate of new vaccinations was also not a top predictor for either wave. This is likely due to two factors. First, by this stage of the pandemic, many of the countries in our dataset had already achieved high coverage for the primary vaccine series, so the rate of new vaccinations was low. Second, the protective effects of vaccination are not immediate.

Along with our findings from the subset of OECD countries (that sequenced larger amounts of genomic data), our results suggest that viral genomics can offer helpful insights in forecasting efforts as to the likely impact of new variants of concern. Furthermore, we found that measures of genetic distance to the previously circulating variants in each country, overall or in the S-gene, were more or similarly informative as the VOC’s selection coefficient. That is, beyond just the competitive advantage of a new VOC, the local genomic landscape and the VOC’s ability to evade immunity are influential in determining a VOC’s success. This differs from the transition from Delta to BA.1, where fewer people had previously been infected, making high transmissibility alone sufficient for the new variant to spread.

As noted, many countries had missing data, and it was challenging to obtain timely and consistent measures of serological features that reflect immunity. This may have contributed to the low importance of serological features in our predictive model. While serology was a key surveillance tool, as the pandemic progressed and population exposure to SARS-CoV-2 increased, serology became a less reliable indicator of recent infection. When most of the population had been exposed at least once, the prevalence of *N* or *S* antibodies rose, making it difficult to detect reinfections through serological data. Furthermore, numbers of reported cases depend on testing policy and reporting inclinations of different jurisdictions, and testing may have at times focused on those most at risk of a severe outcome, travellers, the general public, or on those with other risks. Polymerase Chain Reaction (PCR) testing is expensive, and particularly during (and after) the first omicron wave, many jurisdictions ceased formal testing for COVID-19 unless the result of the test would impact medical treatment for the individual. Even hospitalization and death data suffer from incomparability across jurisdictions, as protocols, reporting requirements and definitions varied. Yet, these data provide our most comprehensive understanding of the pandemic and are the result of many dedicated collection efforts.

Statistical models like the ones we have presented here have limitations. Indeed, there were many limitations to COVID-19 forecasting efforts in general,^30^ including challenges of data quality and highly stochastic underlying transmission processes. Machine learning methods are an attractive solution for modelling these processes without requiring a mechanistic representation of the determinants of wave size, particularly as we can still incorporate our mechanistic understanding into engineering features such as our variables from sequence data. However, these methods are data-intensive. We bolstered our model with multiple time points per country to counteract the challenge of having many more features than countries with sufficient data for prediction. An alternative approach might instead train the model on a single time point from several prior waves. However, as our results revealed, the relative importance of features changed hugely between waves, potentially reflecting large changes in local epidemiological context. It is important to acknowledge that our analysis was conducted retrospectively, and thus is not presented as a real-time forecasting tool. Although we held out all information from the model after the end of our fitting windows, we did not take into account the possibility of reporting delays in the genomic data or other data streams. Rather, we present this work as an exploratory analysis to identify which factors were predictive during the Omicron BA.1 and BA.2 waves. We note, however, that while many predictive efforts are hampered by reporting delays, the genomic features central to our model may be more robust to this challenge and could be valuable in future forecasting frameworks, as they rely on VOC proportions and not total counts.

Determining the relative size of the next variant-driven wave remains an important challenge, both for SARS-CoV-2 and future pandemics. Here we presented a systematic procedure for modelling two VOC waves, providing insights into the driving factors of Omicron BA.1 and BA.2 using the same approach, and yet there are differences in feature importance reflecting the evolving nature of reality and the value of information. As a future VOC emerges, it is unclear to what extent vaccination and previous infection confer immunity, and it is unclear how populations, governments, and public health institutions will respond (and how data will be reported). We sought to determine how well the relative wave sizes of BA.1 and BA.2 could be predicted, and the results were reasonably good, particularly for the question of whether the new wave will be larger or smaller than the most recent one. However, it remains unclear to what extent past waves will be good models for future waves. To date, Omicron variants and recombinants have not evolved to become more severe (as Delta was), though in evolutionary terms there is little to prevent this from occurring. Although there is considerable knowledge of the mutations that govern increased transmissibility and immune escape, with the data currently available, it is challenging to characterize the portion of a given population that will be protected from a new variant. If it is deemed worthwhile to predict the burden and timing of a new variant, additional data on its infectivity and immune evasion in the relevant population will likely be required.

## 5 Data and code availability

All data and code from this study are accessible on GitHub at https://github.com/ShabMolan/VOC. This repository contains accession numbers for all GISAID sequences used in this study, also accessible at doi.org/10.55876/gis8.241023pk.

## 6 Acknowledgements

We gratefully acknowledge all data contributors, including the Authors and their Originating laboratories responsible for obtaining the specimens, and their submitting laboratories for generating the genetic sequence and metadata and sharing via the GISAID Initiative, on which this research is based. We thank the countries who collected and shared their data with the various sources used in this work (OWID, Johns Hopkins University COVID-19 Data Repository, Oxford COVID-19 Government Response Tracker, Google Community Mobility Reports, SeroTracker) as well as the teams that have worked to maintain these websites.

We thank Dr Finlay Maguire, Dalhousie University, for support and technical discussions in the early stages of the project.

## 7 Funding

The authors received funding from the Natural Sciences and Engineering Research Council of Canada https://www.nserc-crsng.gc.ca/index_eng.asp through Discovery Grants RGPIN-2023-4509 (JES) and RGPIN-2019-06624 (CC), and the Canadian Institutes of Health Research https://cihr-irsc.gc.ca/e/193.html through the CGS-M program (SM). The authors were supported by the Federal Government of Canada’s Canada 150 Research Chair programme https://www.canada150.chairs-chaires.gc.ca/home-accueil-eng.aspx (CC), the Canadian Network for Modelling Infectious Diseases (CANMOD) https://canmod.net/ (CC), the Public Health Agency of Canada https://www.canada.ca/en/public-health.html (ML, VG, CLM), and the Genomics Research and Development Initiative (GRDI) by the Government of Canada https://grdi.canada.ca/en (NKS). The funders had no role in study design, data collection and analysis, decision to publish, or preparation of the manuscript.

## Notes

### Competing Interest Statement

The authors have declared no competing interest.

### Funding Statement

The authors received funding from the Natural Sciences and Engineering Research Council of Canada through Discovery Grants RGPIN-2023-4509 and RGPIN-2019-06624 and the Canadian Institutes of Health Research through the CGS-M program. The authors were supported by the Federal Government of Canada Canada 150 Research Chair programme and the Canadian Network for Modelling Infectious Diseases (CANMOD) and the Public Health Agency of Canada and the Genomics Research and Development Initiative (GRDI) by the Government of Canada.

