## supplemental material for "Drivers of COVID-19 variant wave dynamics: inferring oncoming wave size using global data with genomics"

### 1.1 Genomic Feature Equations

#### 1.1.1 Average Genetic Distance

For each pairwise comparison, we can express the genetic distance, $D_i$, between each circulating sequence and the emerging variant in two ways:

1) The raw number of unique substitutions in each circulating sequence, $C_i$, compared to the emerging lineage, $E$:

$$D_i = C_i - (C_i \cap E),$$

or

2) The proportion of each circulating lineage’s substitutions that are unique:

$$\hat{D}_i = 1 - \frac{(C_i \cap E)}{C_i},$$

where $C_i$ is the number of amino acid substitutions relative to wildtype for the $i$th sequence in the fitting window, and $(C_i \cap E)$ is the number of amino acid substitutions shared by the emerging lineage and the $i$th sequence in the fitting window (relative to wildtype).

The mean of all raw genetic distances, $D_i$ (version 1), was used in the random forest model to represent the average genetic distance, $\overline{D}$,

$$\overline{D} = \frac{1}{n}\sum_{i=1}^{n} D_i,$$

where $n$ is the total number of sequences in the fitting window.

#### 1.1.2 Diversity

First, the Shannon Index was calculated using the *vegan* package. The Shannon Index, H, is defined as:

$$H = -\sum_{i=1}^{R} p_i \ln(p_i),$$

where $R$ is the total richness (i.e., number of unique pango lineages) and $p_i$ is the proportion of R for the $i$th lineage.

The Shannon Index was then exponentiated to get the Hill Diversity, H':

$$H' = e^H.$$

### 1.2 Data and missingness

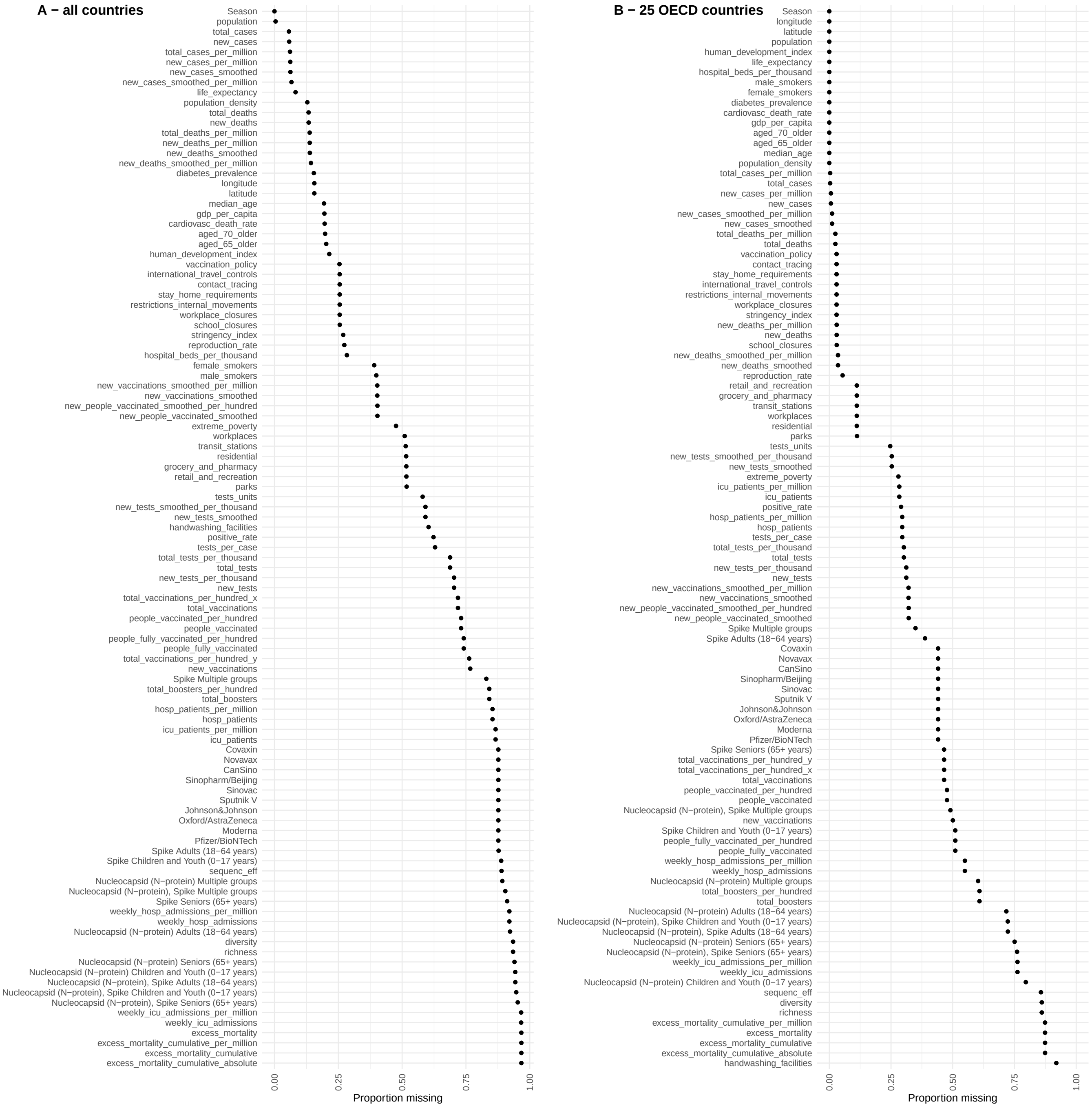


Figure 1: Proportion of variables that are missing, for full time period 2020-01-01 - 2023-02-01. **(A)** Full dataset containing all countries. **(B)** Filtered dataset containing 25 OECD countries only.

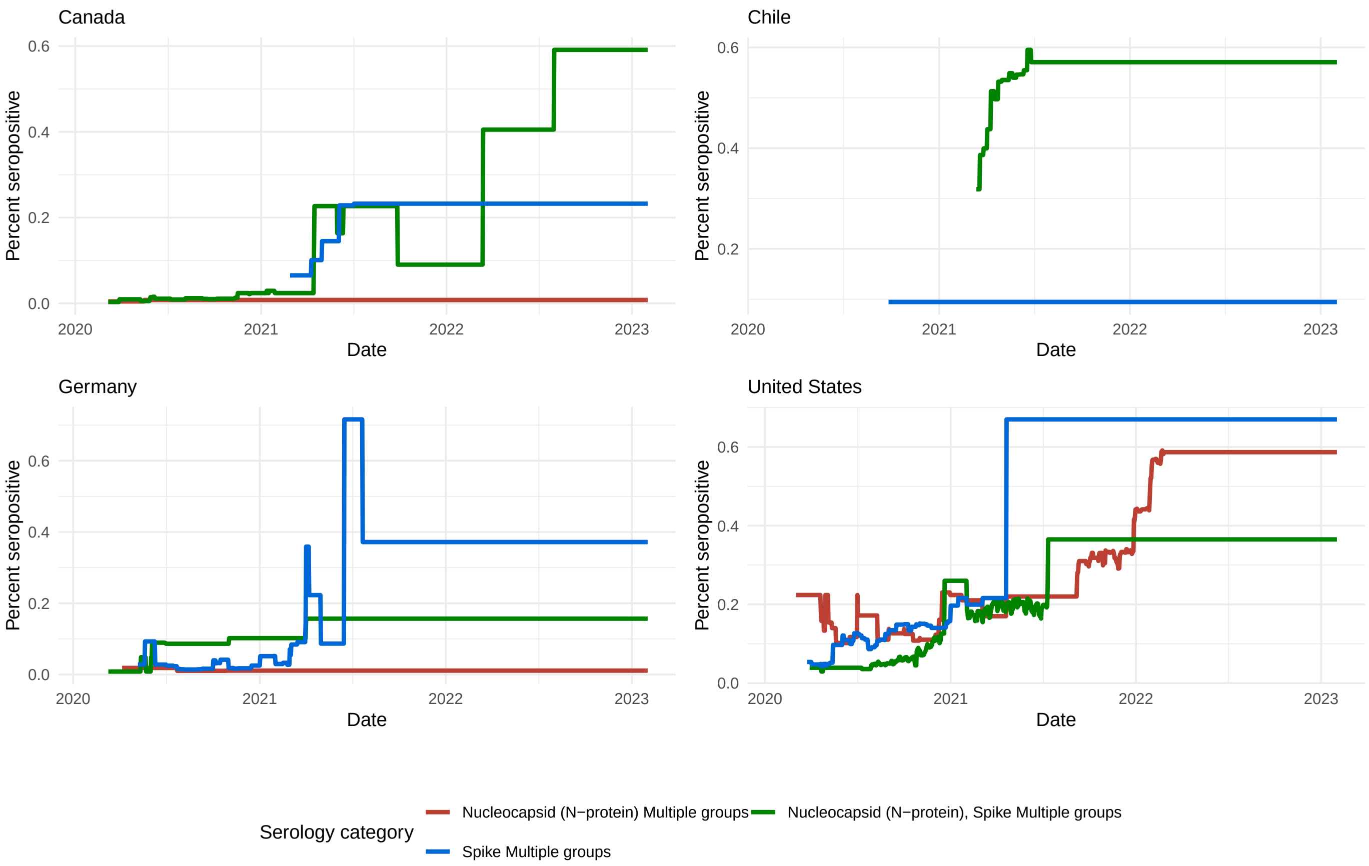


Figure 2: Serology data for a demonstrative selection of OECD countries. Data is plotted after LOCF imputation, to generate piece-wise constant time series of the most recent serology study in each country. Some countries (e.g. Chile) do not have serology data available for all categories (e.g. N-protein).

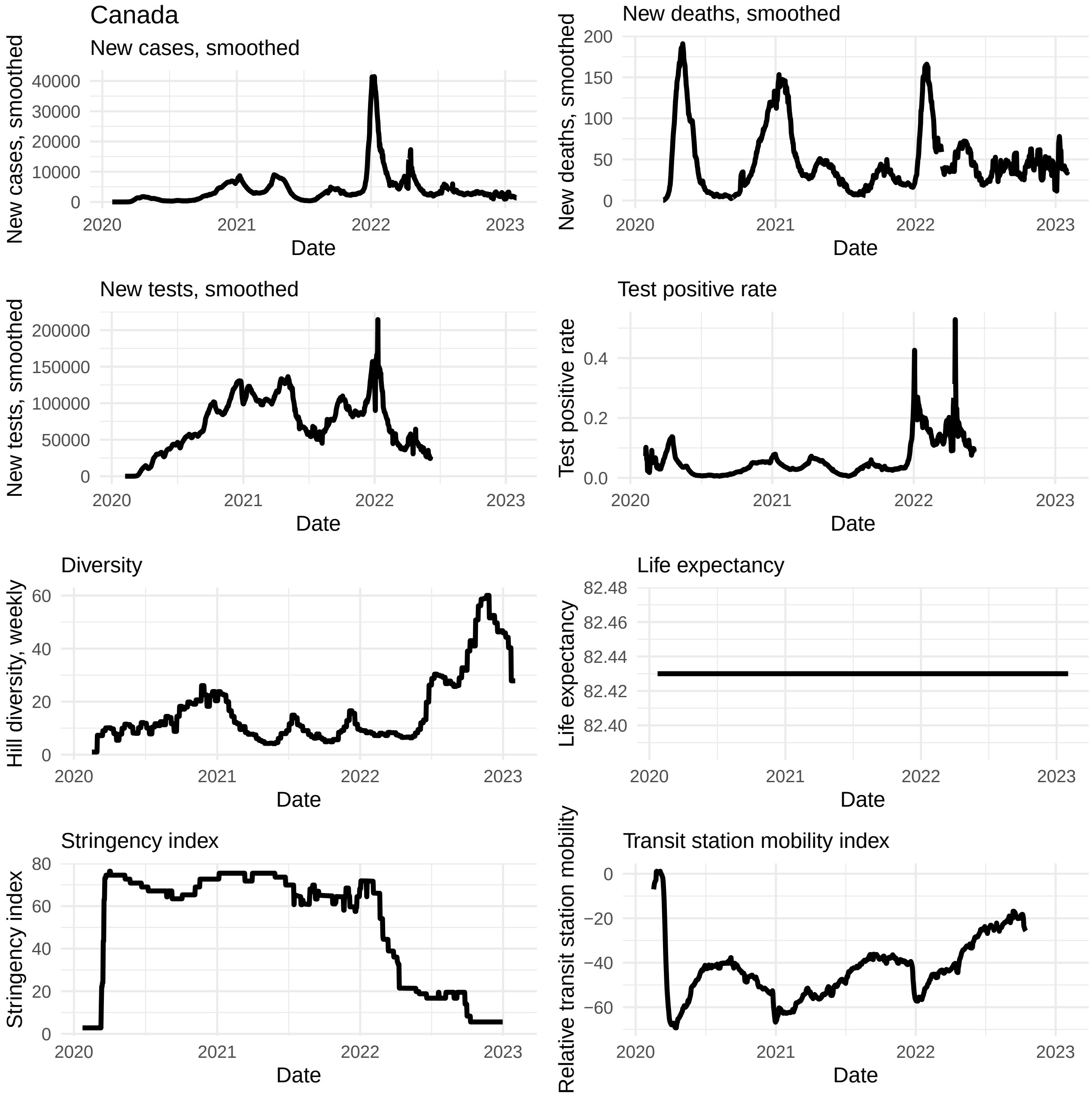


Figure 3: Demonstrative selection of variables from master data set for Canada, for full time period 2020-01-01 - 2023-02-01. Includes variables: smoothed new cases, smoothed new deaths, smoothed new tests (all 7-day smoothed), test positivity rate (rolling 7-day average), Hill diversity (weekly), life expectancy (non-time-varying), government response stringency index (0-100 scale) and Google transit station mobility index (relative to baseline period Jan 3rd - Feb 6th 2020, rolling 7-day average).

### 1.3 Wave dynamics

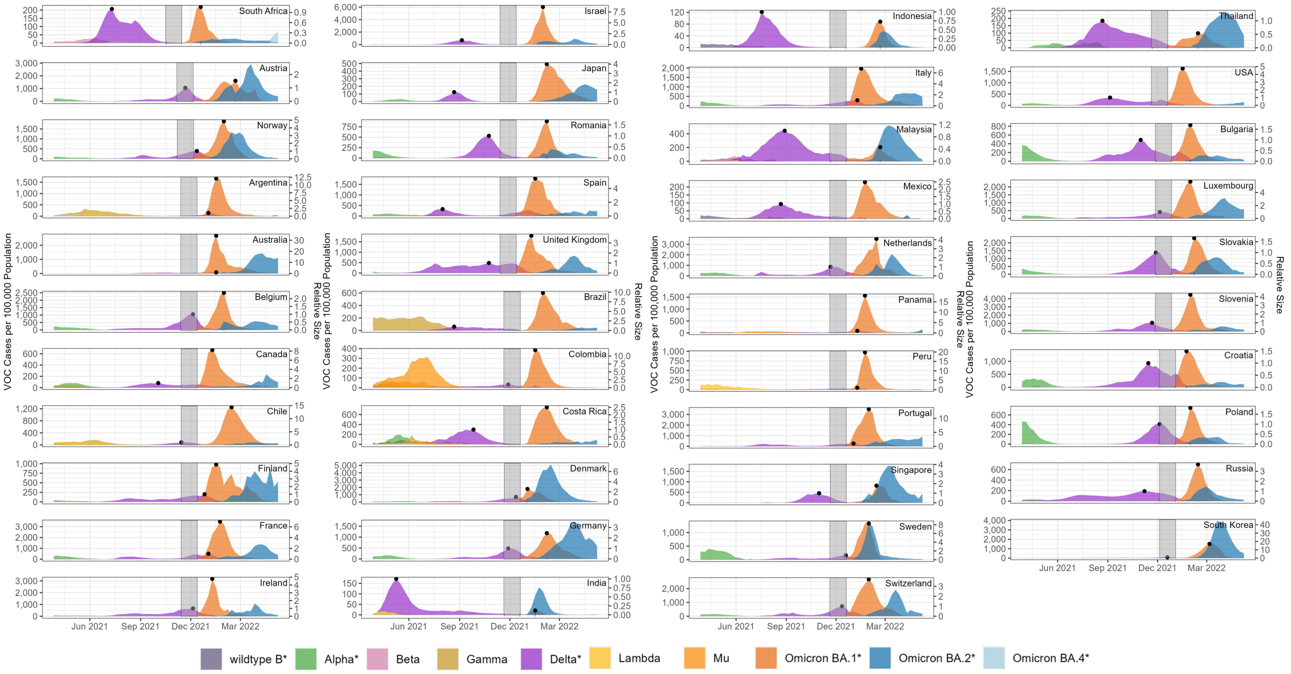


Figure 4: Delta-BA.1 wave dynamics for all 43 countries, columns ordered by the emergence week of BA.1 ($t_0$). $t_0$ tends to occur after the Delta peak, with the exception of Norway, Australia, Finland, France, Argentina, Italy, Panama, Peru, Portugal, and Sweden. Earliest $t_0$ is in South Africa during the week ending on 2021-11-14. Latest $t_0$'s are in Croatia, Poland, Russia, and South Korea during the week ending on 2022-01-02.

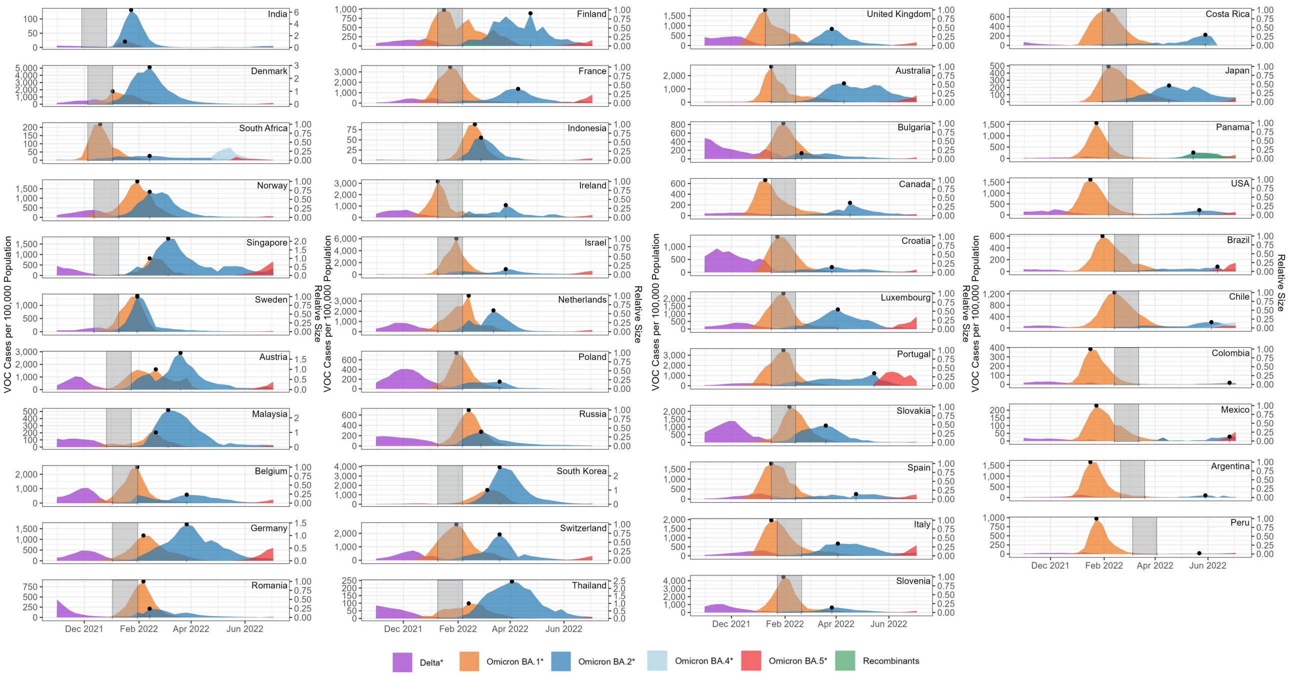


Figure 5: BA.1-BA.2 wave dynamics for all 43 countries. Columns are ordered by the emergence week of BA.2 ($t_0$). Earlier emergence is correlated with a larger BA.2 wave. Later $t_0$ tend to occur after the BA.1 peak, while earlier $t_0$ tend to occur before the BA.1 peak. Earliest $t_0$ is in India during the week ending on 2021-12-26. Latest $t_0$ is in Peru during the week ending on 2022-04-03.

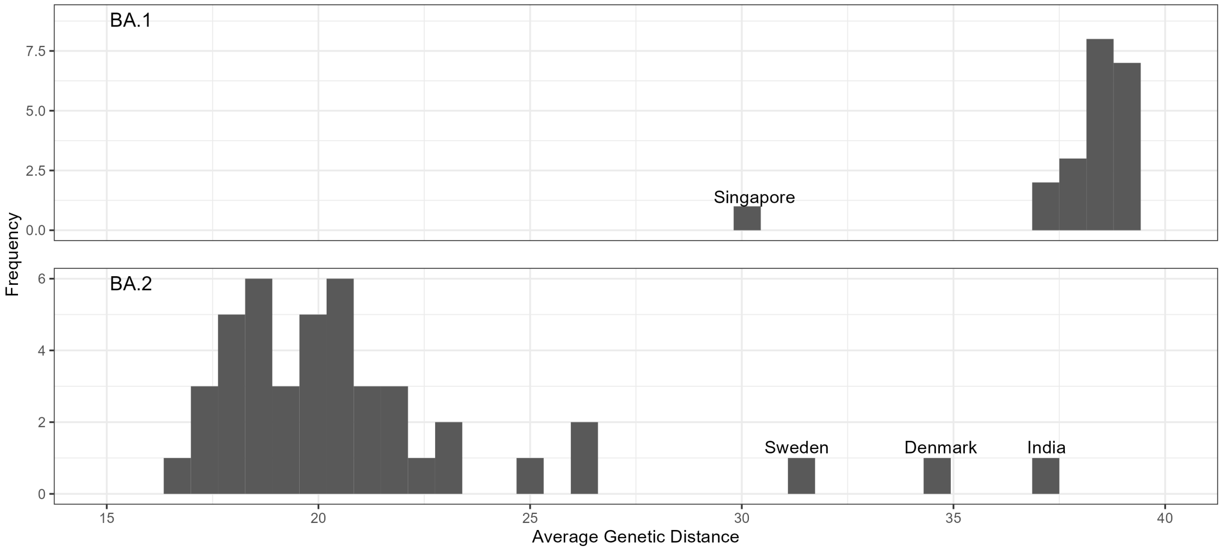


Figure 6: Average Genetic Distance of BA.1 and BA.2 compared to the circulating lineages in their respective fitting windows. Compared to BA.2, BA.1 shows higher genetic distance overall. Within BA.2, the countries where BA.2 first emerged show the highest genetic distance.

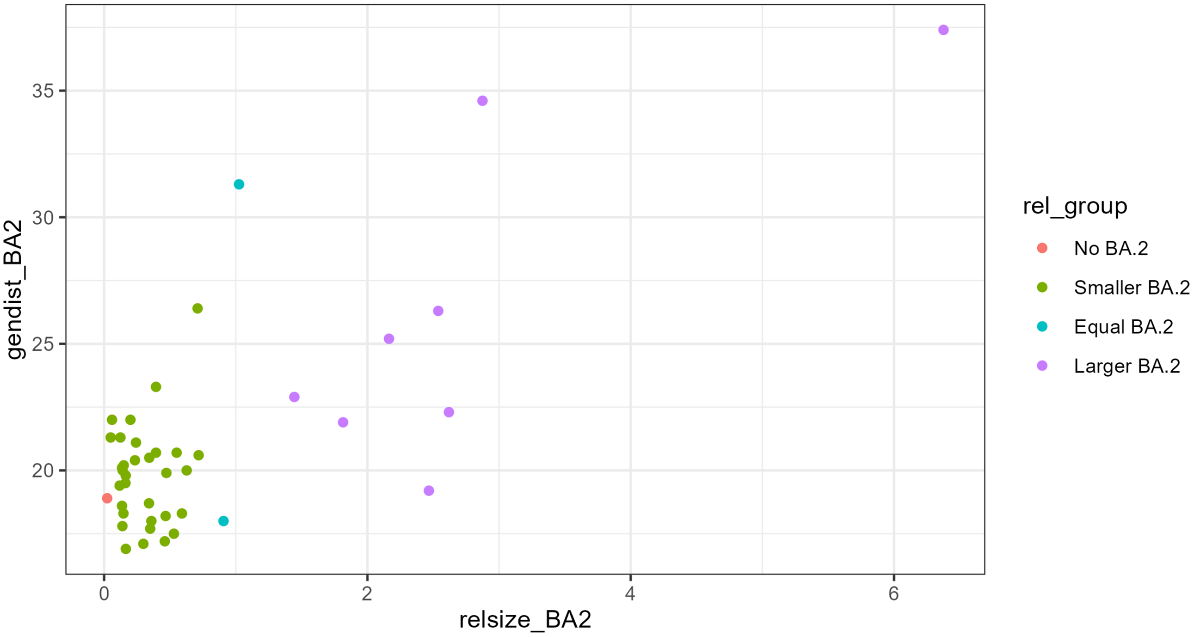


Figure 7: Genetic Distance of BA.2 plotted against the relative size of BA.2 compared to BA.1. There is a general positive trend: as genetic distance increases, so does the relative size of BA.2.

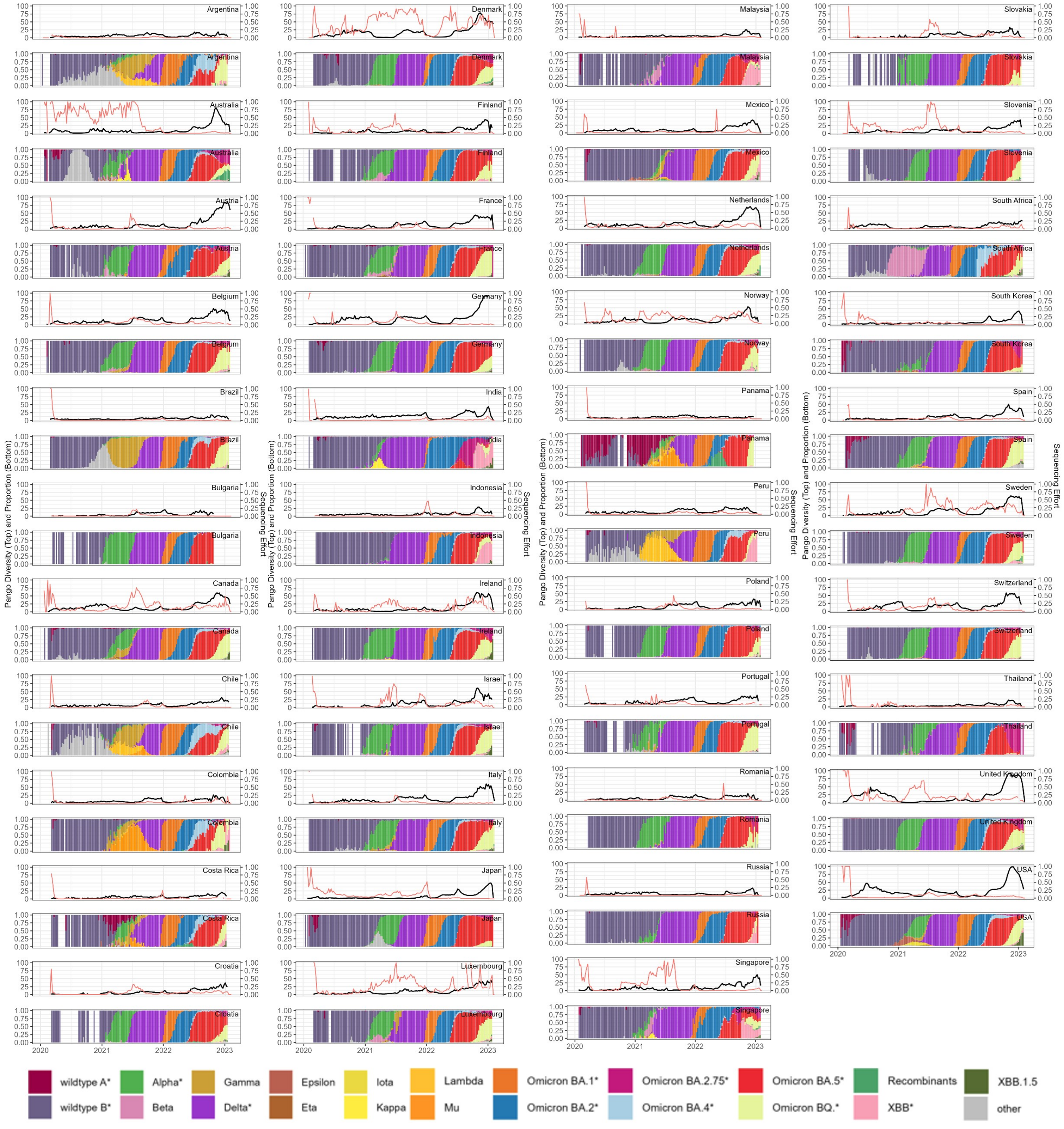


Figure 8: Hill Pango lineage diversity (top of each pair, black line), sequencing effort (top of each pair, red line), and VOC/VOI Proportion (bottom of each pair) for all 43 Countries.

### 1.4 Model predictions for OECD countries

In this section, we present the results of a model trained only on a subset of 25 OECD countries, found to have more robust data with lower missingness (Figure 1).

Panel A of Figure 9 shows the predictions of the random forest model with leave-one-country-out cross validation, with the response variable modelling the relative sizes of the BA.1 to Delta waves. Here, the RMSE of the random forest model is 4.04 and $R^2$ is 0.39. The range of ratios of Delta to BA.1 for OECD countries is between 1.525 and 34.095. Predictions are largely quite good, though Australia is an outlier. In restricting to OECD countries, we observe relatively less uncertainty between models trained on different imputations of the missing data (smaller confidence intervals).

Panel B of Figure 9 shows predictions for the BA.2 to BA.1 relative wave size, again with leave-one-country-out cross validation. The RMSE of the random forest model is 0.37 and $R^2$ is 0.21. The range of ratios of BA.2 to BA.1 for OECD countries is between 0.050 and 2.874. Denmark is the outlier here.

The models correctly predicted whether the subsequent wave is larger, smaller, or equal size in 100% and 88% of the OECD countries for transition from Delta to BA.1 (panel A) and BA.1 to BA.2 (panel B), respectively (Figure 10). Overall, compared to the models for 43 countries, models for OECD countries have smaller RMSE, smaller uncertainty of prediction per country, and higher percentage of correct prediction for the general trend of the subsequent waves.

Figure 11 shows the feature importance plot of the models trained on OECD countries. The top panel ranks features based on importance score calculated as the mean decrease in the mean squared error (MSE) of the BA.2/BA.1 model. The bottom panel shows the scores for the BA.1/Delta model. The most important features for transition between Delta and BA.1 include life expectancy, Human Development Index, S-gene genetic distance, and number of days since the peak of Delta wave. The most important features for transition between BA.1 and BA.2 include genetic distance

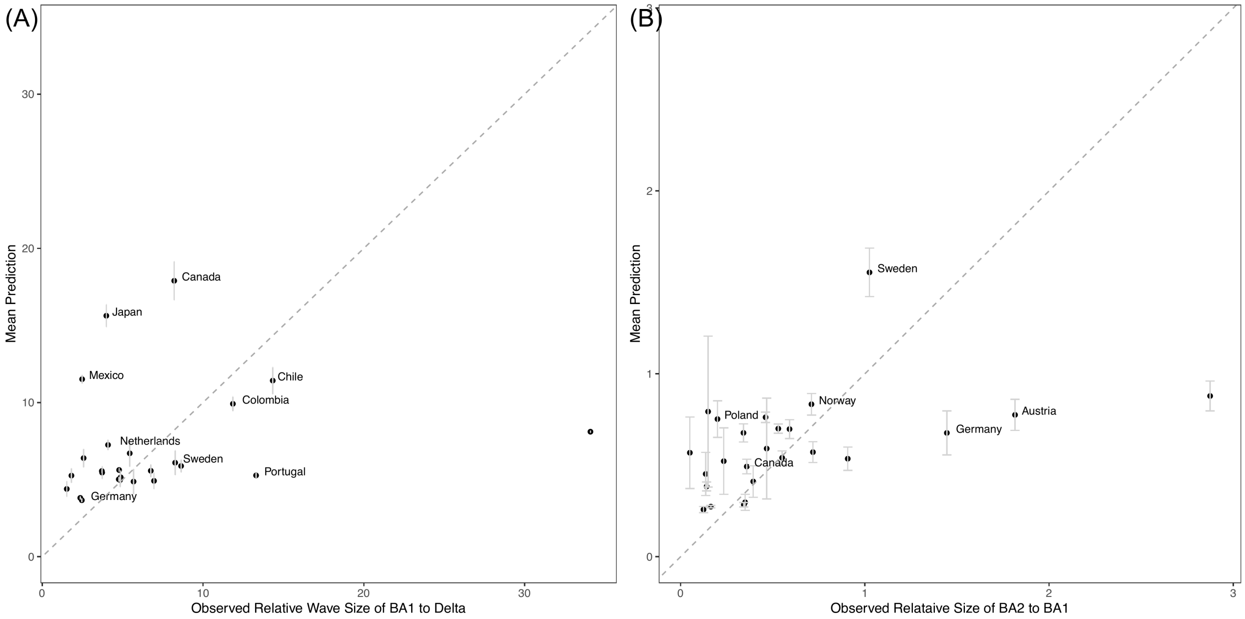


Figure 9: Mean predictions of the relative wave size of the (A) BA.1 to Delta and (B) BA.2 to BA.1 variants across OECD countries using a Leave-One-Country-Out cross-validation approach with a Random Forest model. The x-axis represents the observed relative wave sizes. The y-axis shows the corresponding mean predicted values by the model. The dashed line represents the 1:1 line, indicating perfect agreement between observed and predicted values. Error bars represent the standard deviation of the predictions across multiple imputed datasets.

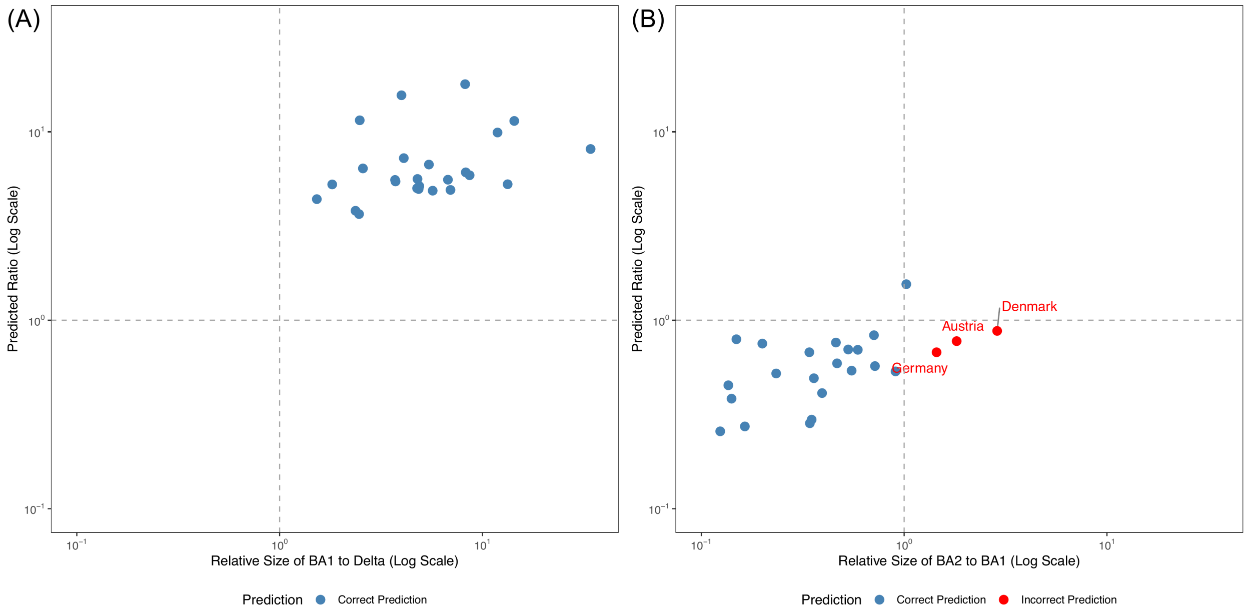


Figure 10: Prediction accuracy plots for two models for OECD countries: (A) BA.1 to Delta and (B) BA.2 to BA.1 variants, both on a log scale. Points in the first and third quadrants represent correct predictions where the model accurately identifies if the ratio of the subsequent waves is larger or smaller than 1. Points in the second and fourth quadrants represent incorrect predictions, where the model incorrectly identifies the ratio of the subsequent waves.

measures (S-gene, total), GDP per capita, and Human Development Index. This again suggests that both the inherent characteristics of the variants themselves and demographic features play key roles in this transition. Note that S-gene genetic distance is an important feature both across different datasets and different wave transitions.

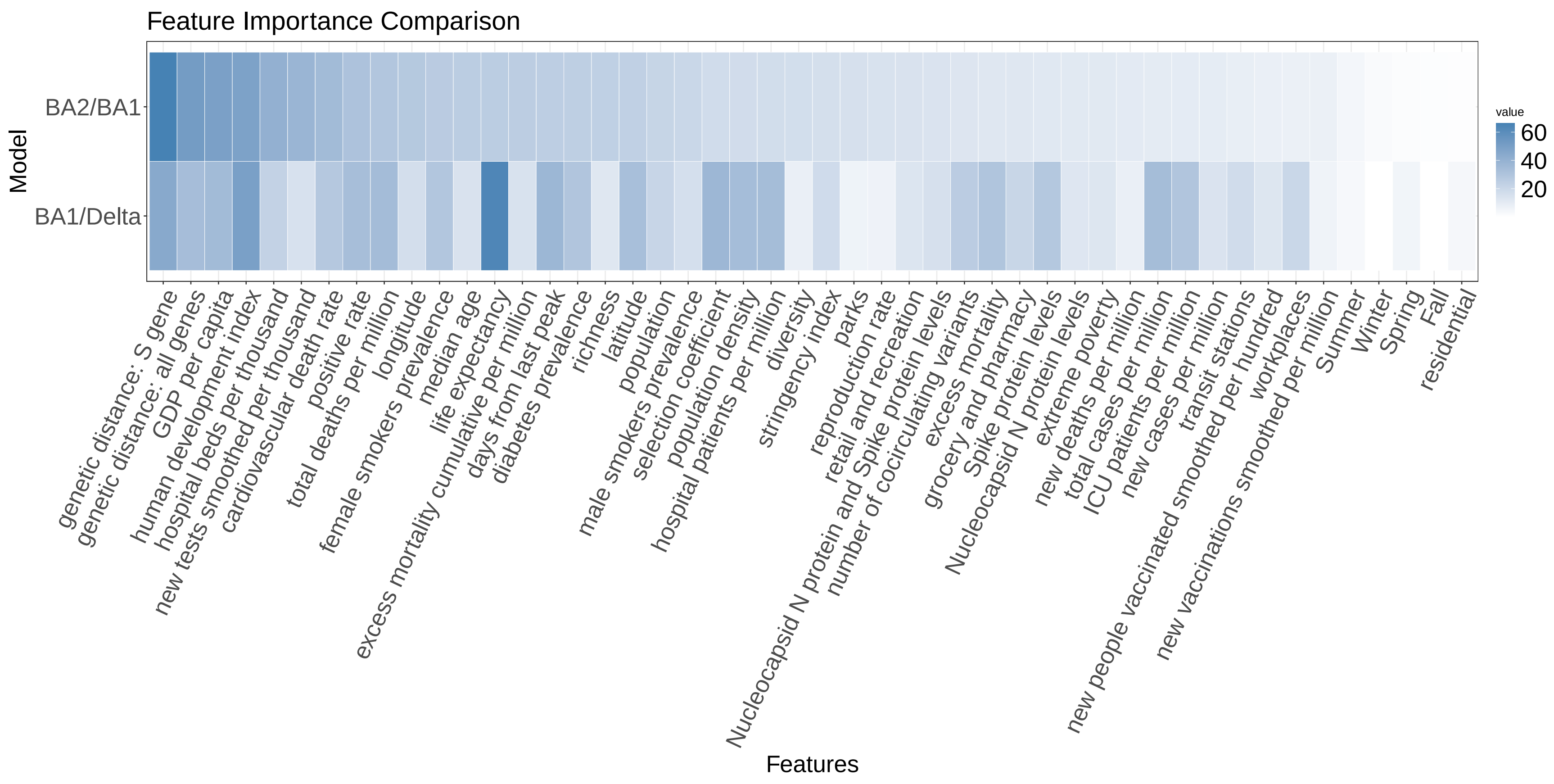


Figure 11: Feature importance comparison for Random Forest models predicting the relative wave size of BA.1 to Delta and BA.2 to BA.1 among OECD countries. The heatmap shows the importance scores of various features for each model, with darker shades indicating higher importance. Feature importance is measured by the percentage increased in mean squared error (MSE) of the model's predictions when a given predictor is removed.

| | |
|---|---|
| 43 countries | Argentina, Australia, Austria, Belgium, Brazil, Bulgaria, Canada, Chile, Colombia, Costa Rica, Croatia, Denmark, Finland, France, Germany, India, Indonesia, Ireland, Israel, Italy, Japan, Luxembourg, Malaysia, Mexico, Netherlands, Norway, Panama, Peru, Poland, Portugal, Romania, Russia, Singapore, Slovakia, Slovenia, South Africa, South Korea, Spain, Sweden, Switzerland, Thailand, United Kingdom, United States |
| OECD countries | Australia, Austria, Belgium, Canada, Chile, Colombia, Denmark, Finland, France, Germany, Ireland, Israel, Italy, Japan, Luxembourg, Mexico, Netherlands, Norway, Poland, Portugal, Spain, Sweden, Switzerland, United Kingdom, United States |

Table 1: List of countries

| Data Category | Features |
|---|---|
| COVID-19 cases and reproduction rates | Total cases per million |
| | New cases per million |
| | Reproduction rate |
| COVID-19 deaths | Total deaths per million |
| | New deaths per million |
| Excess mortality | Excess mortality |
| | Excess mortality cumulative per million |
| COVID-19 hospitalizations and ICU | Hospital patients per million |
| | ICU patients per million |
| Policy responses | Stringency index |
| COVID-19 testing and test positivity | New tests smoothed per thousand |
| | Positive rate |
| COVID-19 vaccination | New vaccinations smoothed per million |
| | New people vaccinated smoothed per hundred |
| COVID-19 serology studies | Nucleocapsid N protein levels |
| | Nucleocapsid N protein and Spike protein levels |
| | Spike protien levels |
| COVID-19 genomic variables | Richness |
| | Diversity |
| | Selection coefficient |
| | Genetic distance: S gene |
| | Genetic distance: all genes |
| | Number of co-circulating variants |
| Demographic | Population |
| | Population density |
| | Median age |
| | Life expectancy |
| | GDP per capita |
| | Human development index |
| | Extreme poverty |
| | Hospital beds per thousand |
| | Cardiovascular death rate |
| | Diabetes prevalence |
| | Male smokers prevalence |
| | Female smokers prevalence |
| Miscellaneous | Residential (Google Mobility) |
| | Workplaces (Google Mobility) |
| | Parks (Google Mobility) |
| | Transit stations (Google Mobility) |
| | Grocery and pharmacy (Google Mobility) |
| | Retail and recreation (Google Mobility) |
| | Latitude |
| | Longitude |
| | Number of days from last peak |
| | Seasons |

Table 2: Features and Data Categories
